# DEUBIQUITINASES AS PROGNOSTIC BIOMARKER AND POTENTIAL DRUG TARGET FOR GYNECOLOGICAL CANCERS

**DOI:** 10.1101/2025.04.22.25326173

**Authors:** Kondapally Mavika, Anubha Dey, Golagana Velangani Harshitha, Shashi Kiran, Manjari Kiran

## Abstract

**Objective:** To develop Deubiquitinase-Associated Signatures (DAS) to predict the prognosis of gynecological cancer patients.

**Method:** Using a cox-lasso regression model, we have developed Deubiquitinase-associated signatures for Cervical, Ovarian, and Uterine cancers. Developed DAS were validated in TCGA and GEO datasets. Survival analysis was carried out to know the effect of factors like menopausal stage and grade on DAS. The survival prediction accuracy of DAS was analyzed using ROC curves. Immune infiltration scores of 22 immune subtypes were explored using the CIBERSORT package in risk groups classified by DAS. Further, to target the unfavorable deubiquitinases (DUBs), compounds were identified using CMap database.

**Results:** Three DAS were developed for Cervical, Ovarian, and Uterine cancer types. DAS was able to predict survival and classify patients into two groups in TCGA and GEO datasets. DAS is an independent predictor of survival irrespective of tumor grade and menopausal stage. DAS, along with the clinical features, improves the accuracy of predictions. CIBERSORT analysis has shown that Immune cell infiltration is associated with risk groups divided by DAS. Using CMap, 52 compounds were identified to target unfavorable DUBs.

**Conclusion:** DAS is a good predictor of survival, and targeting unfavorable DUBs can decrease tumor progression in gynecological cancers.

**Synopsis:**

- Deubiquitinases (DUBs) are associated with cancer progression, limited studies on gynecological cancers.
- We used cox-lasso regression to develop a DUB-Associated prognostic Signature (DAS).
- DAS stratifies patients into high-low-risk and improves survival prediction accuracy.
- Small molecules were identified that can target poor prognostic DUBs.

## INTRODUCTION

Gynecological cancers (Cervical, ovarian, and uterine) constitute 14.4% of all new cancers affecting women, with significant mortality rates^1^. Identifying outcome predictors in such cancers has been a major effort of researchers around the world to better manage such patients ^2^. Gene expression signatures of the tumors that can distinguish long-surviving patients from short-surviving ones can be used for differentiating high and low-risk patients and their prospective management. Prognostic gene signatures based on analysis of all genes have been identified for cervical^3–5^, ovarian^6,7^, and uterine cancers^8,9^. In cancers, including gynecological cancers, ubiquitination is the most commonly altered pathway in cancers^10^. Ubiquitination is carried out by a large group of genes many of which are involved in cancers^11^. Accordingly, ubiquitination based prognostic genes were identified in cervical ^12–14^, ovarian ^15–17^ and uterine cancers^18^. While ubiquitination is a very large family of proteins with ∼600 genes, deubiquitination is carried out by a small family of ∼100 deubiquitinases or DUBs. Unlike ubiquitination protein complexes, which are multiprotein complexes, DUBs are single or few-unit enzyme proteins with a distinct enzymatic pocket that can be targeted for therapy^11^. Therefore, DUBs can be targeted for manipulation of the ubiquitination process in cancers. A DUB-based signature for Hepato-cellular carcinoma (HCC) was recently reported, which also identified possible inhibitors that can be used for the treatment of such cancers^19^. However, no attempt has been reported to identify DUBs of prognostic significance in gynecological cancers. Therefore, we used the TCGA expression data to identify prognostic DUB signatures in cervical, ovarian, and uterine cancers that effectively distinguish patients with better survival from poor ones. We also characterized the immune infiltration of high-risk patients identified by our DUB signature and identified inhibitors that can target DUBs with positive/ negative survival coefficients.

## MATERIALS AND METHODS

### 1. Data Collection

Gene expression data (HTseq-FPKM) and clinical information of the patients were downloaded from the UCSC XENA (https://xenabrowser.net/) for three types of Gynecological cancers (Cervical, Ovarian, and Uterine). Independent test datasets were downloaded from GEO (GSE44001, GSE63885, GSE119041). We downloaded the list of human deubiquitinases from iUUCD(https://iuucd.biocuckoo.org/). The tumor samples with incomplete clinical data were removed from the TCGA RNASeq data. The GEO dataset was processed using the following steps: 1) Samples with incomplete clinical data were removed; 2) the most sensitive probe for each gene was considered. The sample information of 3 TCGA datasets and 3 GEO datasets for Cervical, Ovarian, and Uterine types of cancers are shown in Table S1.

### 2. Construction of Prognostic signature based on Deubiquitinases

Firstly, TCGA gene expression data was z-transformed. Then we divided the TCGA patients into 80% training set and the remaining 20% into unseen-testing set using “caret” package. As various clinical variables can have an effect on the survival of patients, we considered confounding factors for each cancer type. The 80% training set of data was again divided into 70% training and 30% test. We performed multivariate cox regression analysis using the “survival” package on the training set of data, which was used to analyze the prognostic potential of each DUB by controlling the effect of the confounding factors ^20^. The genes with significant association (p<0.05) with prognosis were considered for further steps. LASSO regression analysis was performed using the “glmnet” package. The risk score was calculated for the patients in both the test and train dataset using the signature obtained after cox-lasso analysis. Survcutpoint function in R was used to stratify patients into high-risk and low-risk in both test and training sets. Survival differences between high-risk and low-risk groups were compared using the likelihood ratio test. All the steps mentioned above were repeated 1000 times. We selected the models that showed significant differences in test and training sets. We considered the median risk score of the DUBs from the significant models as the final risk coefficient. Based on the number of times the significant models picked up a particular DUB in 1000 bootstraps, the top 10 DUBs were selected. Among the top 10 DUBs, we selected the DUBs with median cox coefficient >0.15 and variance<0.15 as the final prognostic DUBs.

### 3. Validation of DAS in Training and Independent Datasets

We validated DAS developed for Cervical, Ovarian, and Uterine cancers in unseen 20% testing (TCGA) and independent (GEO) datasets. The risk score was calculated, and the optimal cutoff point was decided using *survcutpoint* function. The cutoff was used to stratify patients into high-risk and low-risk groups. We performed a Kaplan-Meier analysis to estimate the survival rate of patients using the “survminer” package. Log-rank test was performed to compare the survival distributions of the two groups.

### 4. Effect of other factors on DAS

To check if the risk score generated from DAS has an effect by menopause stage and tumor grade, complete TCGA data was divided based on menopausal status and tumor grade, respectively, and the risk score was calculated. As the information on menopausal status was not available for ovarian cancer, we considered the average age of menopause, i.e., (50) as the cutpoint. As mentioned, *survcutpoint* was used to decide the cutoff for stratifying patients. KM plots for Pre and Post menopause-stage patients were plotted, and log-rank test was performed. A similar analysis was performed on patients with different tumor grades for cervical, ovarian, and uterine cancer types.

### 5. Prediction of survival by DAS and other clinical factors

The prediction accuracy of survival by DAS and other clinical characteristics was assessed by comparing the AUC values generated using the pROC package.

### 6. Prediction of Substrate Proteins

After developing the DAS and validating it in independent datasets, we further wanted to target the unfavorable DUBs because the higher the expression of these DUBs, the poorer the prognosis of patients. Firstly, the substrates of DUBs with positive risk coefficient (unfavorable DUBs) for the prognosis of patients in cervical, ovarian, and uterine cancers were identified using UbiBrowser (http://ubibrowser.ncpsb.org). We considered all the experimentally validated known substrates and predicted substrates with a confidence score >0.85 for these DUBs. The network of the DUBs and substrates was visualized using Cytoscape (version 3.10.1).

### 7. Functional Enrichment Analysis

Functional enrichment analysis was performed on the substrates of unfavorable DUBs using “clusterProfiler” package.

### 8. Tumor Immune Microenvironment Analysis

After stratifying the TCGA patients into high-risk and low-risk groups based on DAS, the “ESTIMATE” package was used to calculate the immune, stromal, and estimate scores. To analyze the relation between the immune infiltration of 22 immune cells and two risk groups, “CIBERSOFT” package was used.

### 9. Prediction of Drug sensitivity and drug screening

In-vivo drug responses of each sample in two risk groups divided using DAS were predicted using the “OncoPredict” package based on GDSC data(https://www.cancerrxgene.org/) for the commonly used chemotherapy agents for cervical, ovarian, and uterine cancer types.

A connectivity map (https://clue.io/) database was used to identify small-molecule drugs that target unfavorable DUBs.

## RESULTS

### 1. Deubiquitinase-associated signatures (DAS) for cervical, ovarian, and uterine cancers

The RNASeq data for all three gynecological cancers was downloaded from UCSC Xena, which was then split into 80:20 train and test datasets. Cox-lasso regression was performed on the training set of data to identify DUBs associated with survival [<u>Figure S1</u>].

After training the cox-lasso model on 80% of the TCGA dataset, the top 10 DUBs were selected. Among the top 10 DUBs, the DUBs with a median cox coefficient greater than 0.15 were considered [<u>Figure S2</u>]. Finally, three deubiquitinase-associated signatures (DAS) were developed for Cervical, Ovarian, and Uterine cancer types. The signatures developed for three cancer types are as follows:

**Uterine Cancer**

(0.260*OTUD7A) + (0.174*USP26) + (-0.162*OTUD7B) + (0.155*STAMBP) + (0.164*COPS5)

**Ovarian Cancer**

(-0.1513*USP18) + (-0.1587*USP28) + (-0.1759*USP51) + (0.1511*OTUD7A) + (0.1521*USP43)

+ (-0.1714*USP6)

**Cervical Cancer**

(0.2147*USP12) + (-0.1878*OTUD7A) + (-0.2453*MPND) + (0.1796*UFSP1) + (-0.1571*UFD1L) + (0.1561*OTUB2)

Risk scores calculated for each DAS are in the following format.

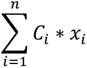

Where, 𝐶*_i_* is the risk coefficient, which was calculated for each DUB by the cox-lasso model, and 𝑥*_i_* is the expression value of each DUB.

In the signature, the DUBs with positive risk coefficients act as unfavorable DUBs for prognosis, whereas the DUBs with negative risk coefficients act as favorable DUBs for prognosis. OTUD7A gene was a common DUB identified as prognostic and part of the signature in all three cancers. However, the role of OTUD7A was different; it acts as an unfavorable DUB in ovarian and uterine cancers and as a favorable DUB in cervical cancer.

### 2. DAS is a predictor of survival in the training and independent dataset

To evaluate the performance of DAS in survival prediction, we calculated the risk score using DAS for each patient in unseen TCGA (20%), TCGA complete, and GEO datasets. The patients were stratified into high-risk and low-risk based on the optimal cutoff generated using *survcutpoin*t. Survival analysis has shown that the high-risk group has poorer survival rates than the low-risk group, except in the uterine external validation set [<u>Figure S3</u>]. The patients can be significantly divided into high-risk and low-risk, indicating that the DAS can predict the survival of patients with cervical, ovarian, and uterine cancers [Figure 1]. The expression of prognostic DUBs in both risk groups is shown in <u>Figure S4</u>.

**Figure 1:**
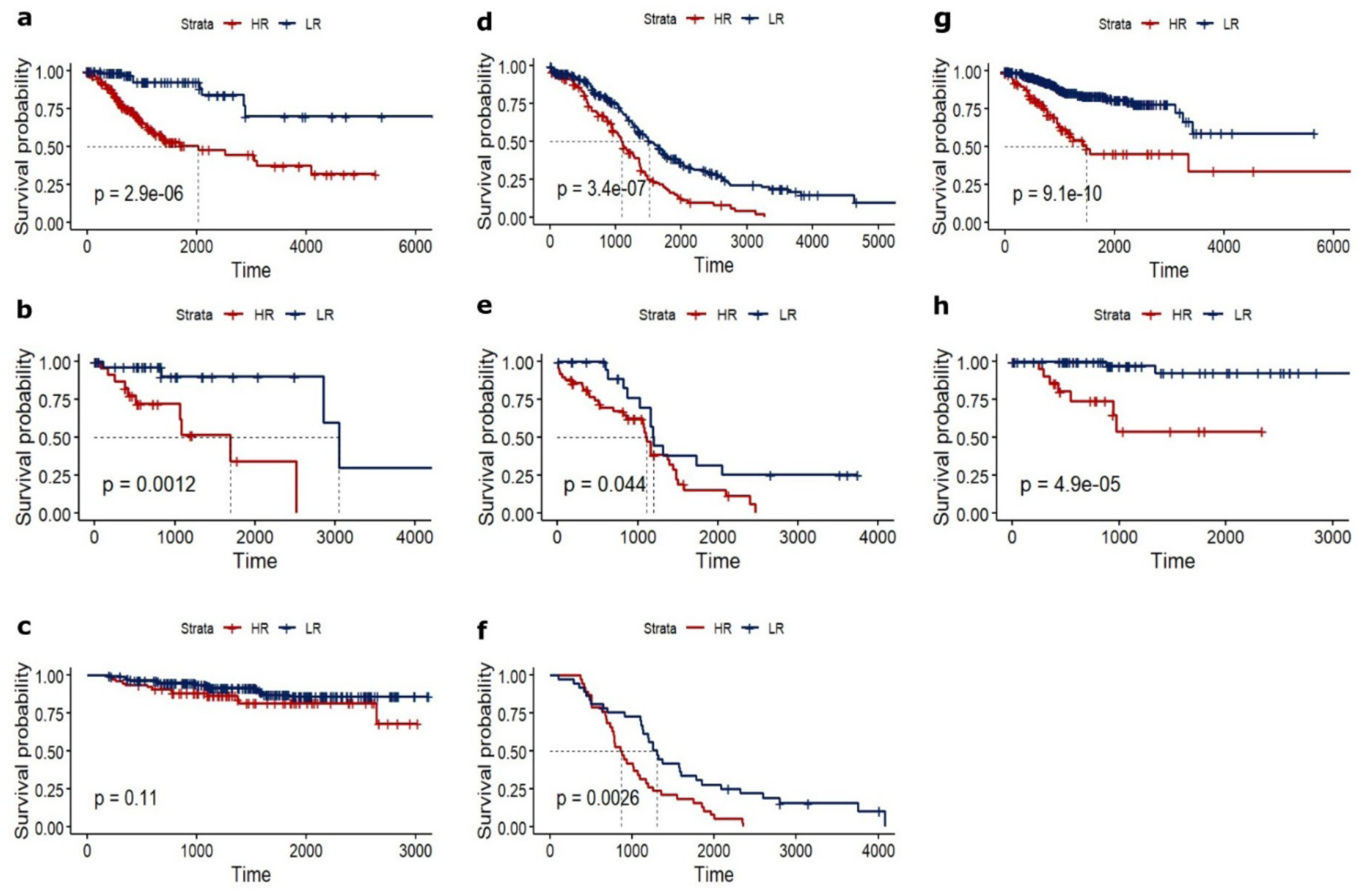
Validation of DAS in TCGA complete, TCGA test, and GEO datasets in Cervical (a,b,c), Ovarian(d,e,f) cancers and validation in TCGA complete, TCGA test in Uterine (g,h) cancer respectively. DAS, Deubiquitinase associated signatures; TCGA, The Cancer Genome Atlas; GEO, Gene Expression Omnibus

### 3. DAS is an independent predictor of survival

To check if DAS can predict high-risk and low-risk groups in different grades and menopausal stages, the dataset was divided. Patients in the postmenopausal stage were higher in ovarian and uterine cancer and less in cervical cancer [Figure 2a-c]. In the same way, patients with high-grade cancer were more in ovarian and uterine cancer dataset and less in cervical cancer [Figure 2j-l]. KM curves have shown that DAS significantly stratified the patients into two groups. KM plots of patients with differences in menopause status and grades are shown in Figure 2. This analysis shows that DAS can categorize patients into high-risk and low-risk patients irrespective of their menopausal stage [Figure 2d-i] and tumor grade [Figure 2m-t]. This indicates that DAS is an independent predictor of survival, and there is no effect of menopausal stage and tumor grade on DAS.

**Figure 2:**
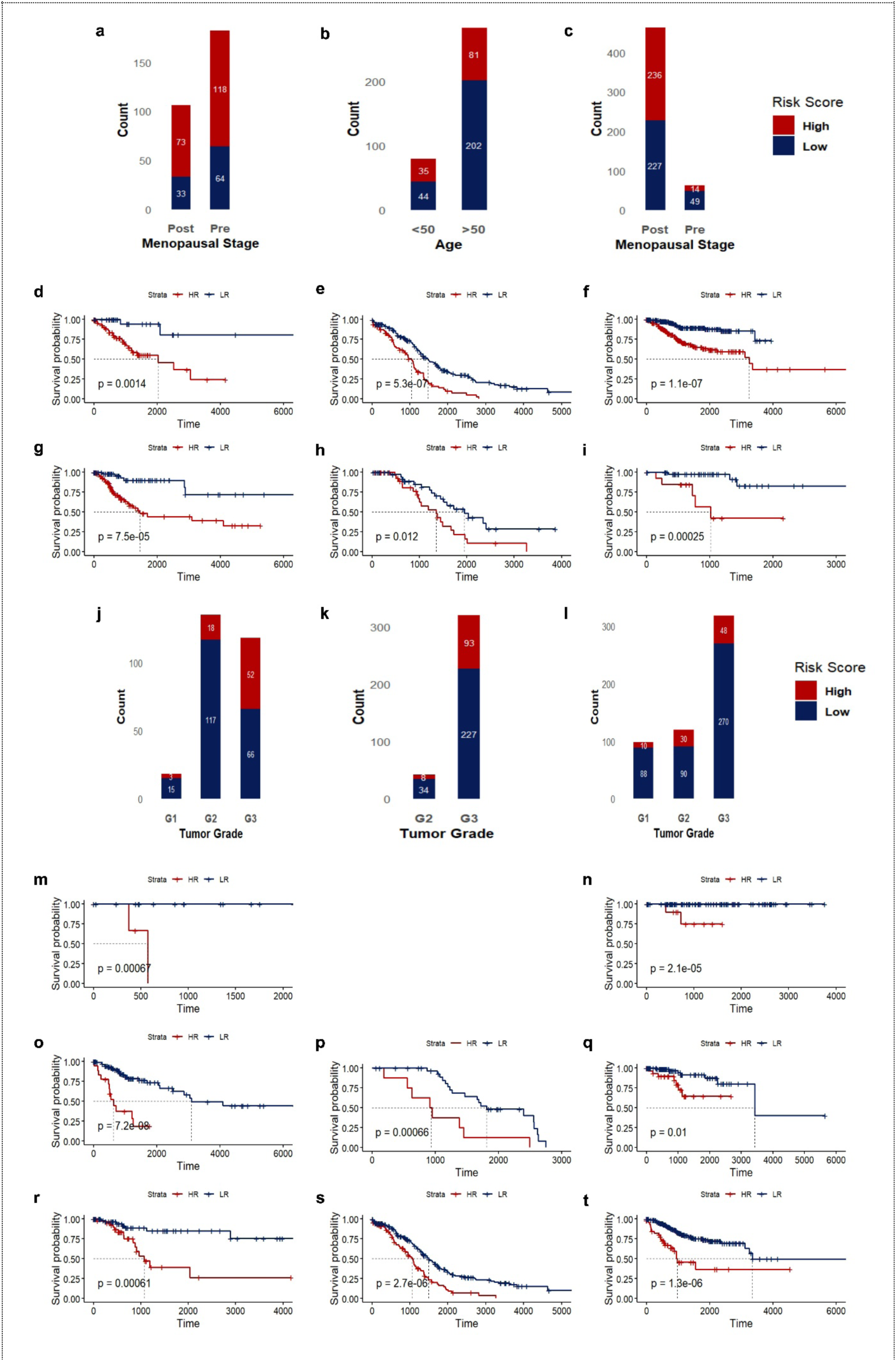
Barplots showing the count based on DAS across the menopausal stage(a,b,c), tumor grade(j,k,l), and Survival analysis of two risk groups of patients in post-menopause(d,e,f),pre-menopause (g,h,i), G2 (o,p,q), G3 (r,s,t) in Cervical, Ovarian and Uterine cancers respectively and G1 (m,n) in Cervical and Uterine cancers. DAS, Deubiquitinase associated signatures

### 4. DAS with clinical variables improves the survival prediction

Clinical factors influence the prognosis of gynecological cancers, and studies have shown that clinical factors act as prognostic factors^21^. However, clinical factors alone cannot be good prognostic markers^22^. Integrating clinical factors with biomarkers can increase the accuracy of survival prediction. To further confer this, we assessed the accuracy of survival prediction by individual clinical variables, a combination of clinical variables, DAS alone, and DAS along with clinical variables (combination), and compared the AUC values. We have observed that DAS, if considered along with clinical variables(combination) improves the survival prediction. It showed the highest accuracy (0.95,0.76,1 in cervical, ovarian, and uterine cancers, respectively) in predicting the patient’s survival compared to individual clinical variable or considering all the clinical variables together [Figure 3]. DeLong test was performed to assess whether the combination model significantly outperformed other models. The combination model has shown significantly higher AUC than other models. This analysis indicates that integrating DAS and clinical variables improves prediction accuracy in all three gynecological cancers.

**Figure 3:**
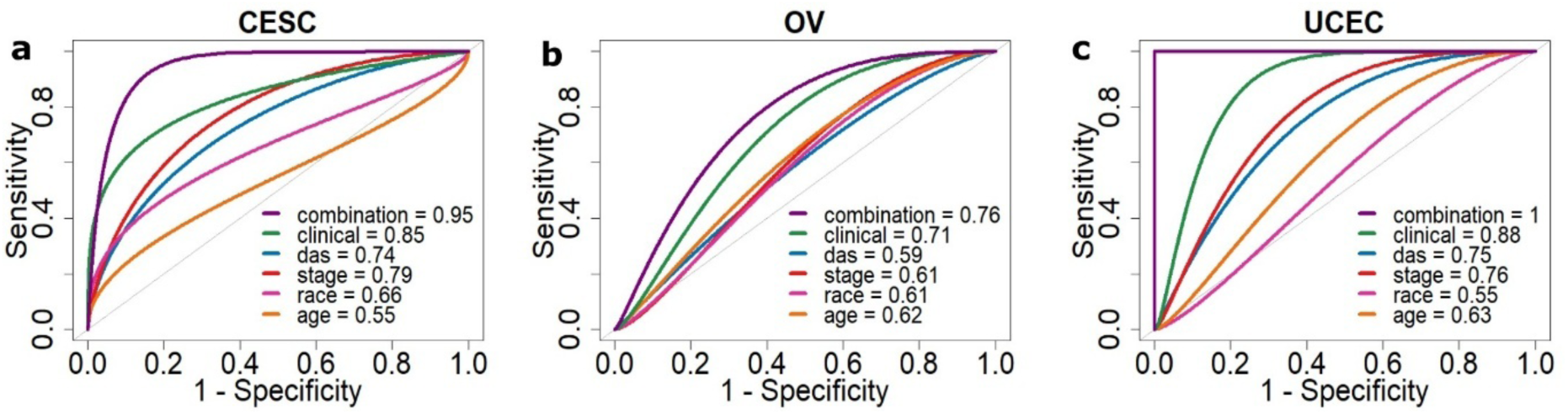
Combined model showing higher AUC compared to clinical factors, DAS, stage, race, and age across Cervical, Ovarian, and Uterine cancers (p < 0.05, DeLong test) DAS, Deubiquitinase associated signatures; AUC, Area under the curve

### 5. Substrates of Unfavorable DUBs

As the expression of unfavorable DUBs leads to poor prognosis, we next identified the drugs that can reduce the expression of target genes. We identified seven DUBs with positive coefficients and, therefore, are poor prognostic DUBs in DAS of cervical, ovarian, and uterine cancer types. UFSP1, an unfavorable DUB in cervical cancer, has no predicted and known substrates, and for the remaining DUBs, 145 substrates were found using UbiBrowser. The network of DUBs and their substrates identifies DUBs with common targets and DUBs targeting multiple proteins [Figure 4a].

**Figure 4:**
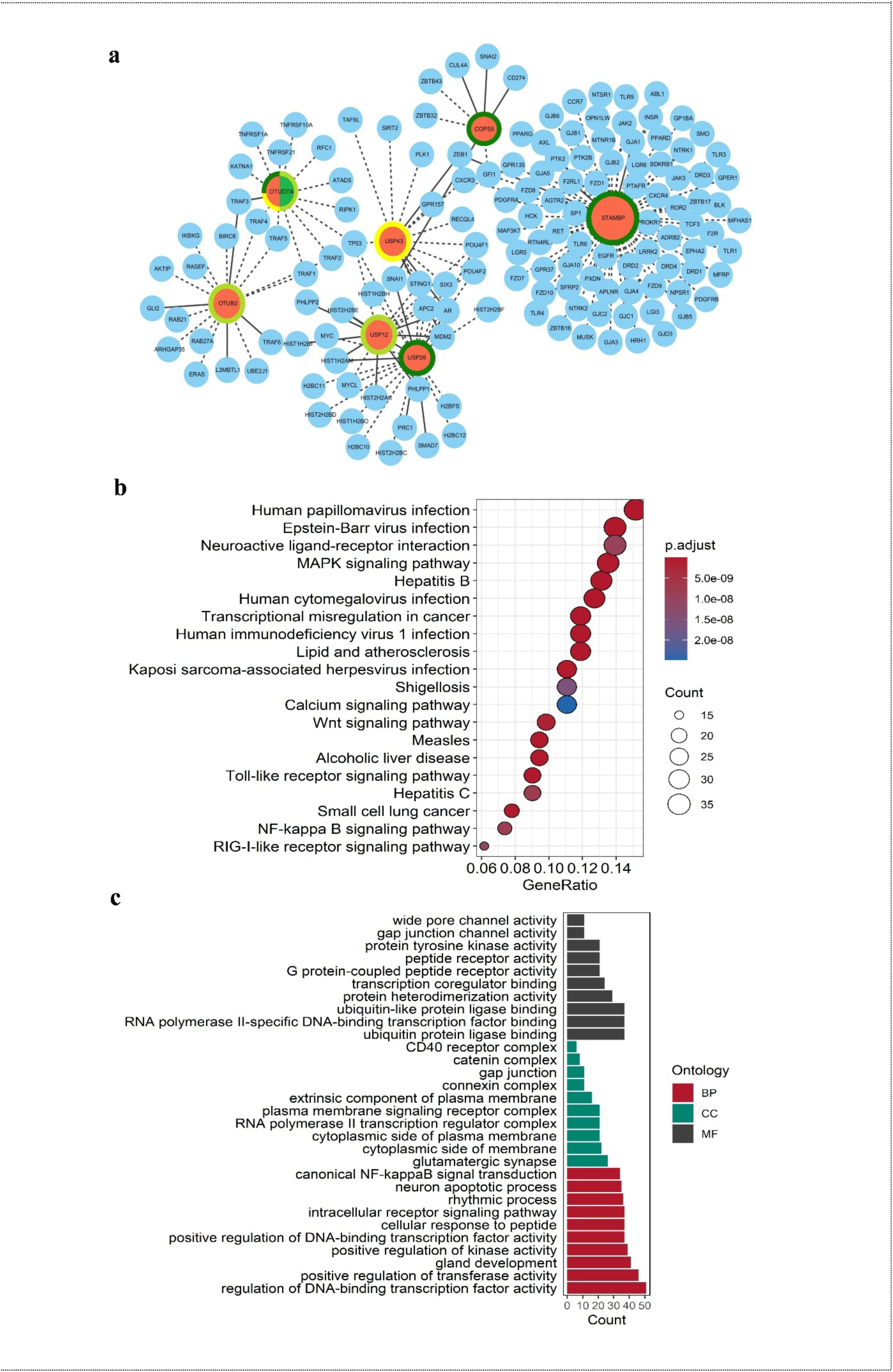
(a)Network of unfavorable DUBs and their substrates. Colored rings around the node.Green: cervical, yellow: ovarian, and dark green: uterine cancer. Dashed lines: predicted substrates, solid lines: known substrates. Size of the node represents degree. (b) KEGG pathway analysis and (c) GO enrichment performed on unfavorable DUB substrates. DUB, Deubiquitinase; KEGG, Kyoto Encyclopedia of Genes and Genomes; GO, Gene Ontology.

Interestingly, OTUD7A, belonging to the OTU family of DUBs, was a common DUB present in the signatures of all three gynecological cancers considered in the present study. It acts as a favorable marker in cervical cancer and unfavorable in the case of ovarian and uterine cancer. Studies suggest that OTUD7A exhibits a dual role in tumor suppression^23^ and tumor progression^24^, but the role of OTUD7A in gynecological cancers has never been studied.

### 6. Substrates of unfavorable DUBs associated with gynecological cancer-related pathways

We next sought to find functional enrichment of targets of unfavorable DUBs. Gene set enrichment analysis of substrates of unfavorable DUBs suggests that genes in DAS are involved in various viral infection pathways like Human Papillomavirus Infection, Epstein-Barr Virus Infection, Hepatitis C, Human Immunodeficiency Virus 1 Infection, Kaposi Sarcoma-Associated Herpesvirus Infection and various signaling pathways like MAPK Signaling Pathway, Wnt Signaling Pathway, Calcium Signaling Pathway, NF-kappaB Signaling Pathway, and Toll-Like Receptor Signaling Pathway [Figure 4b]. GO enrichment analysis shows a strong connection between ubiquitin signaling, NF-kappaB activation, and transcription regulation, all influenced by deubiquitinases (DUBs). Given their role in tumor progression, DUBs may be potential regulators and therapeutic targets in gynecological cancers [Figure 4c].

### 7. Immune infiltration

We performed ESTIMATE analysis to explore the differences in the immune response between high-risk and low-risk groups and compared immune cell infiltration. ESTIMATE analysis showed that the low-risk group had elevated immune scores compared to the high-risk in all three gynecological cancers. The stromal and estimate scores were higher in the low-risk group in uterine and cervical cancers [<u>Figure S4</u>]. As the immune scores were higher in the low-risk group, we further evaluated the infiltration of 22 immune cells to identify cell types contributing to effective immune response in low-risk groups. We have observed that the infiltration of immune cells was higher in the low-risk group than in the high-risk group for cervical and uterine cancer, suggesting a stronger anti-tumor immune response and better prognosis^25,26^. This reflects active immune surveillance, a favorable tumor microenvironment that can effectively detect and combat tumor cells [Figure 5a and 5b]. Whereas in the case of ovarian cancer, infiltration of CD4 T cells, CD4 memory resting cells, M0 macrophages, and resting dendritic cells are higher in the high-risk group indicating a weaker, less effective immune response^27^ and the higher levels of dendritic cells and activated NK cells in low-risk suggest a stronger immune response^28,29^ [Figure 5c].

**Figure 5:**
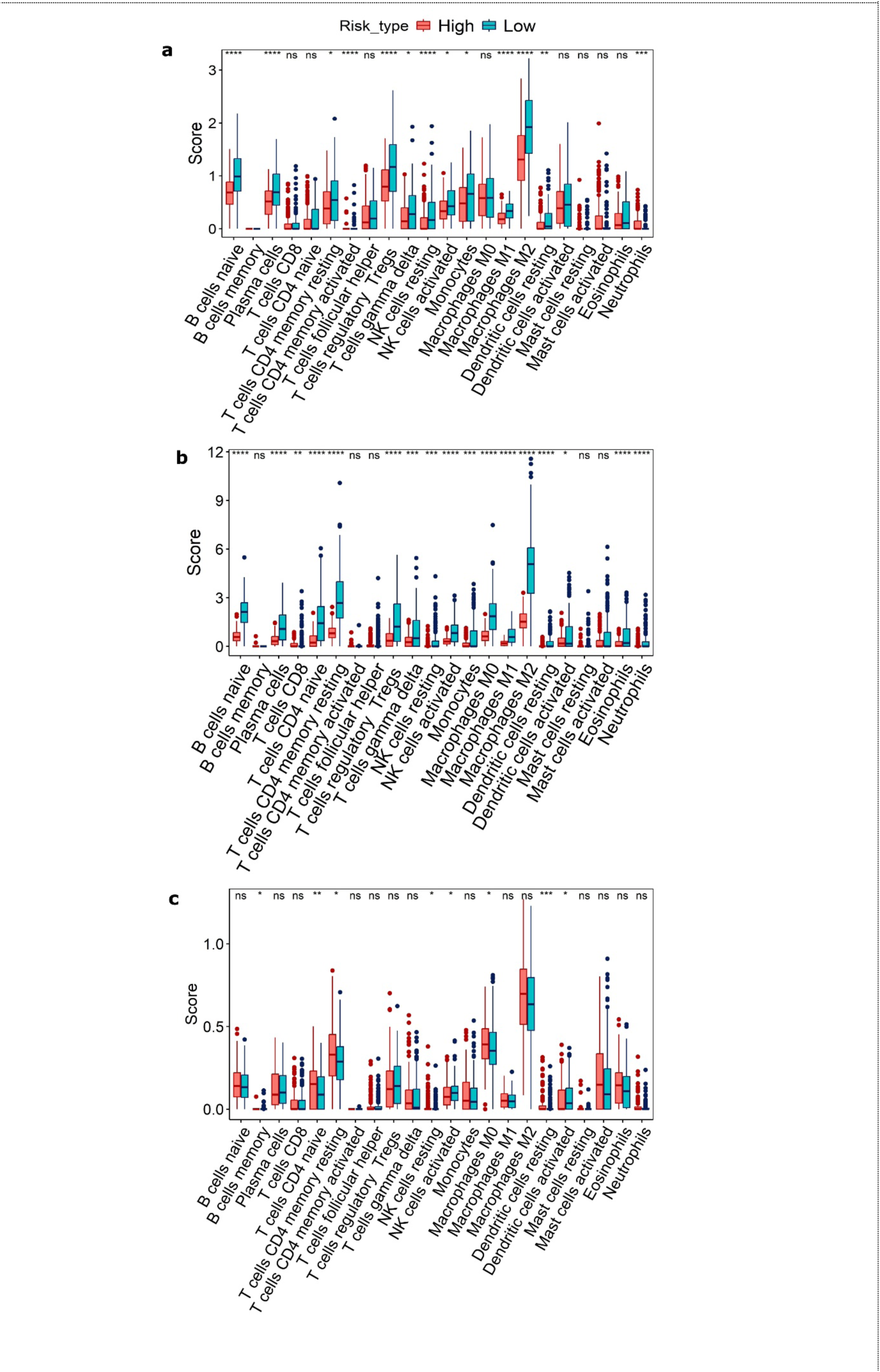
Boxplots showing the difference in the infiltration of immune cells in two risk groups divided using DAS in Cervical(a), Uterine(b), and Ovarian cancers(c). ^ns^ p >0.05, * p<0.05, ** p<0.01, *** p<0.001, **** p<0.0001, wilcox test.

### 8. High-risk group corresponds with low drug sensitivity

To analyze the relationship between DAS and drug sensitivity, we calculated the half-maximal inhibitory concentration (IC50) values of drugs used in chemotherapy of cervical^30^, ovarian^31,32^, and uterine cancers. We observed that the high-risk group showed significantly low drug sensitivity (high IC50) [Figure 6]. These results indicated that the high-risk group patients classified using DAS are associated with resistance to drugs used.

**Figure 6:**
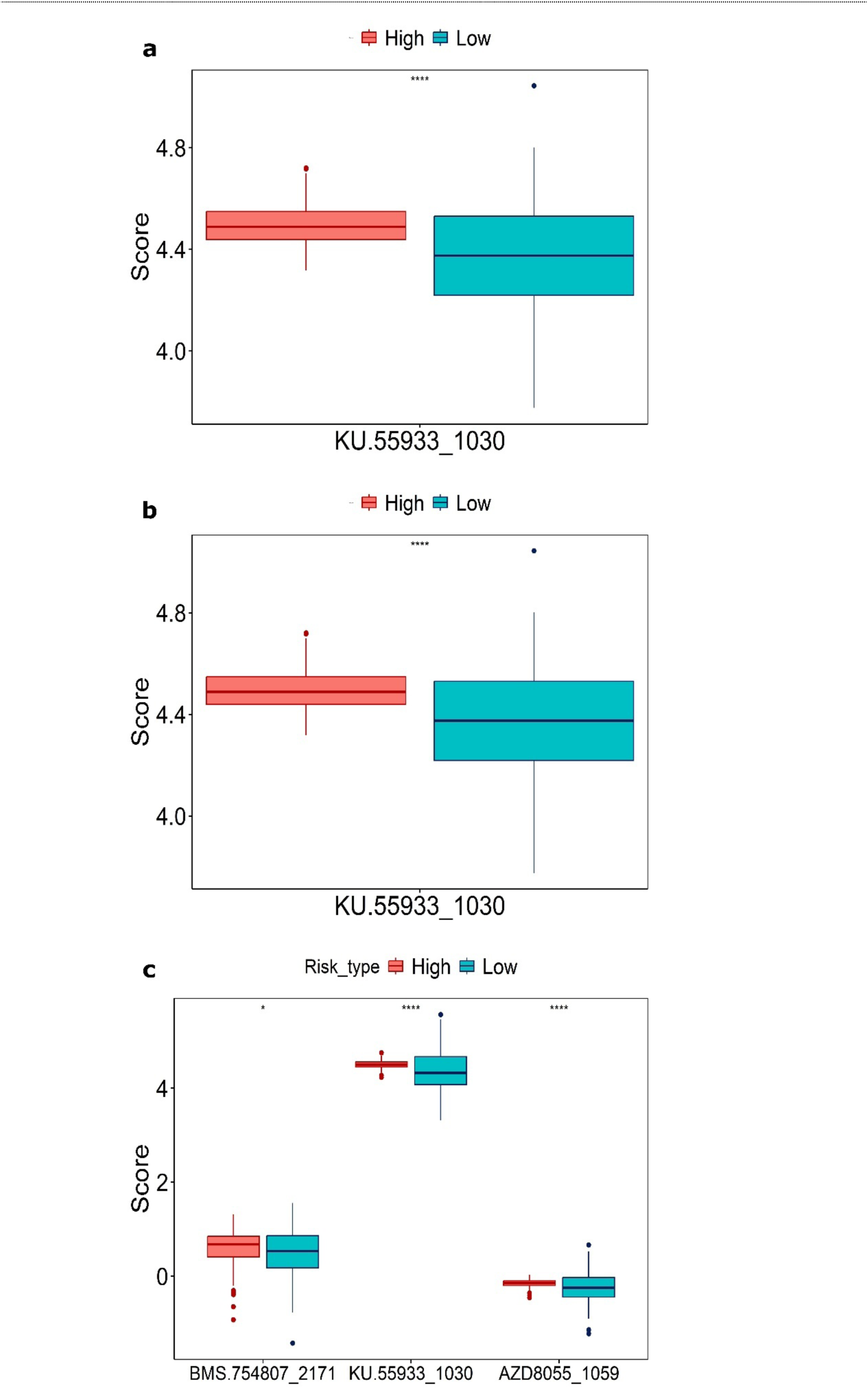
Boxplots showing high drug IC50 in high-risk group in cervical(a), uterine (b), and ovarian (c) cancers. ^ns^ p >0.05, * p<0.05, ** p<0.01, *** p<0.001, **** p<0.0001, wilcox test.

### 9. Small molecular compounds for unfavorable prognostic DUBs

Using the CMap database we identified small molecular compounds to target unfavorable DUBs. As the expression of unfavorable DUBs affects patient’s survival, targeting them can be effective in controlling the tumor progression. We considered the compounds with connectivity score >90 for each DUB [<u>Figure S7</u>]. Few of the compounds identified have potential roles in the treatment of gynecological cancers, such as doxorubicin, capecitabine, pembrolizumab, and metformin.

## DISCUSSION

Gynecological cancers are the most common cancer type in women. Detection of the cancer at the late stage is one of the causes of the high mortality rate worldwide. So, there is a need for biomarkers that can detect the cancer, predict the survival, and can be targeted for efficient treatment. DUBs play a role in tumorigenesis and are emerging drug targets.

In this study, we wanted to investigate the role of DUBs as prognostic marker and built three deubiquitinase- associated signatures for the three top most occurring gynecological cancer types (cervical, ovarian and uterine cancers) using the Cox-Lasso model. The DAS developed effectively predicted the survival of patients in the unseen internal TCGA dataset for the three gynecological cancers and the external GEO dataset for ovarian and cervical cancers. In the case of uterine cancer, the DAS could not predict the survival of risk groups, which was due to the expression difference of three prognostic DUBs, OTUD7A, OTUD7B, and USP26 [<u>Figure S6</u>].

DAS is an independent predictor of survival, irrespective of factors that are associated with the risk of gynecological cancers such as menopausal stage^33^.DAS significantly increases the accuracy of prediction when integrated with clinical factors. Pathway analysis has shown that the substrates of unfavorable DUBs are associated with various infection pathways^34^ and signalling pathways^35,36^ associated with gynecological cancers. Through CIBERSOFT analysis we observed that Immune cell infiltration is associated with the risk score calculated using DAS. Studies have shown the unfavorable DUBs we obtained in our DAS have a role in the tumor progression of that particular cancer. For example, USP12 and OTUB2 are two unfavorable DUBs, in the DAS we constructed for cervical cancer, USP12 regulates the growth of cervical cancer by increasing BMI-1, c-Myc, and cyclin D2 levels^37^. OTUB2 promotes cervical cancer by stabilizing FOXM1^38^, RBM15-mediated m6A modification, and AKT/mTOR signaling^39^. USP43, an unfavorable DUB for ovarian cancer, promotes the growth of ovarian cancer through stabilization of HDAC2 and activation of the Wnt/β-catenin signaling pathway^40^. The studies above support that the expression of unfavorable DUBs in DAS promotes tumor growth. These DUBs can act as new targets in Cervical, Ovarian, and Uterine cancer treatment.

## Data Availability

All codes used are available on https://github.com/Mavikakondapally/codes

## Acknowledgments

We thank the members of SKlab and MKlab for the helpful discussions. KM is a registered Integrated Systems Biology master’s student, AD and GVH are registered PhD students at the University of Hyderabad. AD and MK also acknowledge funding support from UoH-IoE Grant (UoH-IoE-RC2–21–012) and DBT-BUILDER. GVH and SK acknowledge funding support from ICMR (IIRPIG-2024-01-03223) and UoH- IoE Grant (UoH-IoE-RC2-21-020).

---

**Figure S1:**
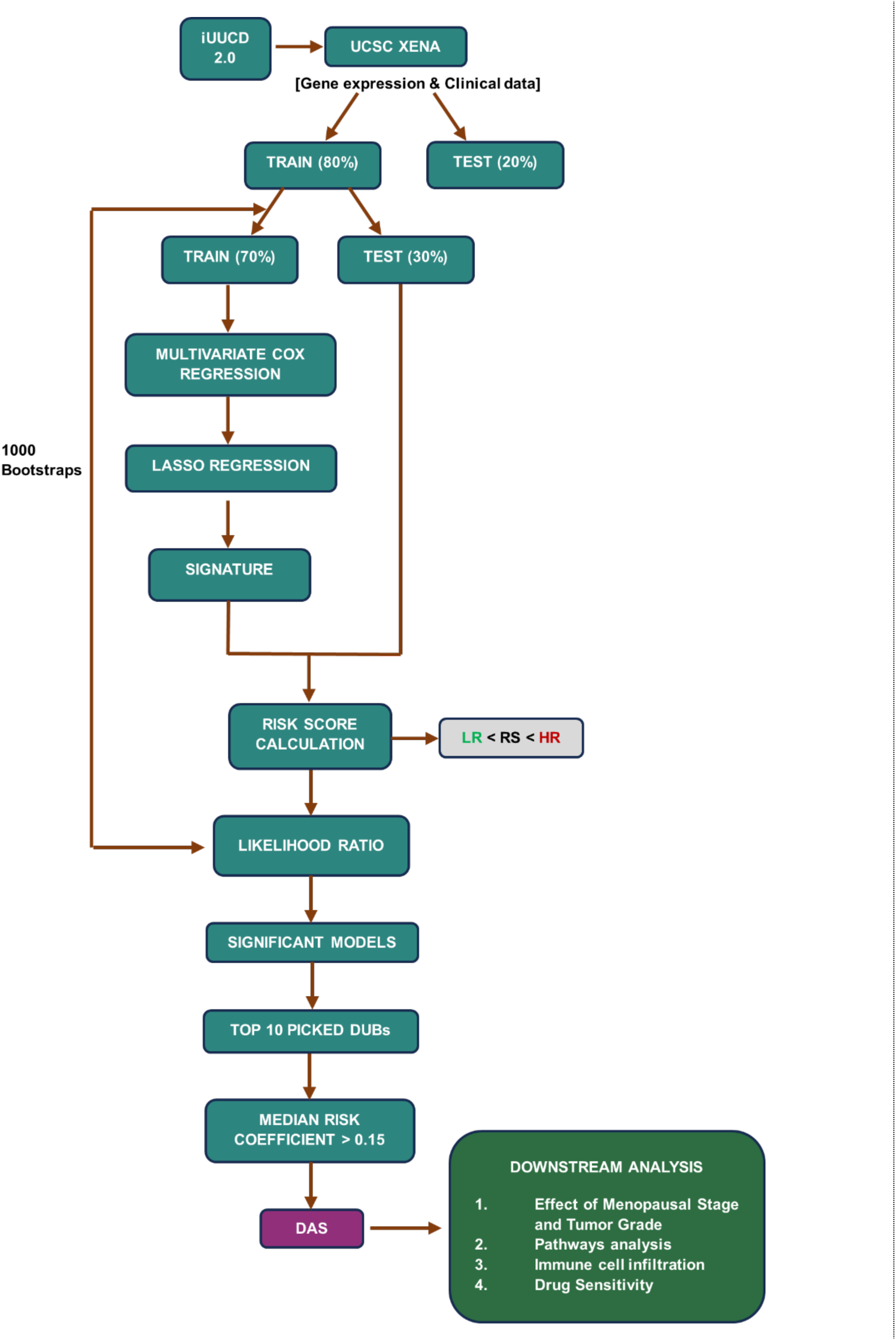
Flowchart of the study

**Figure S2:**
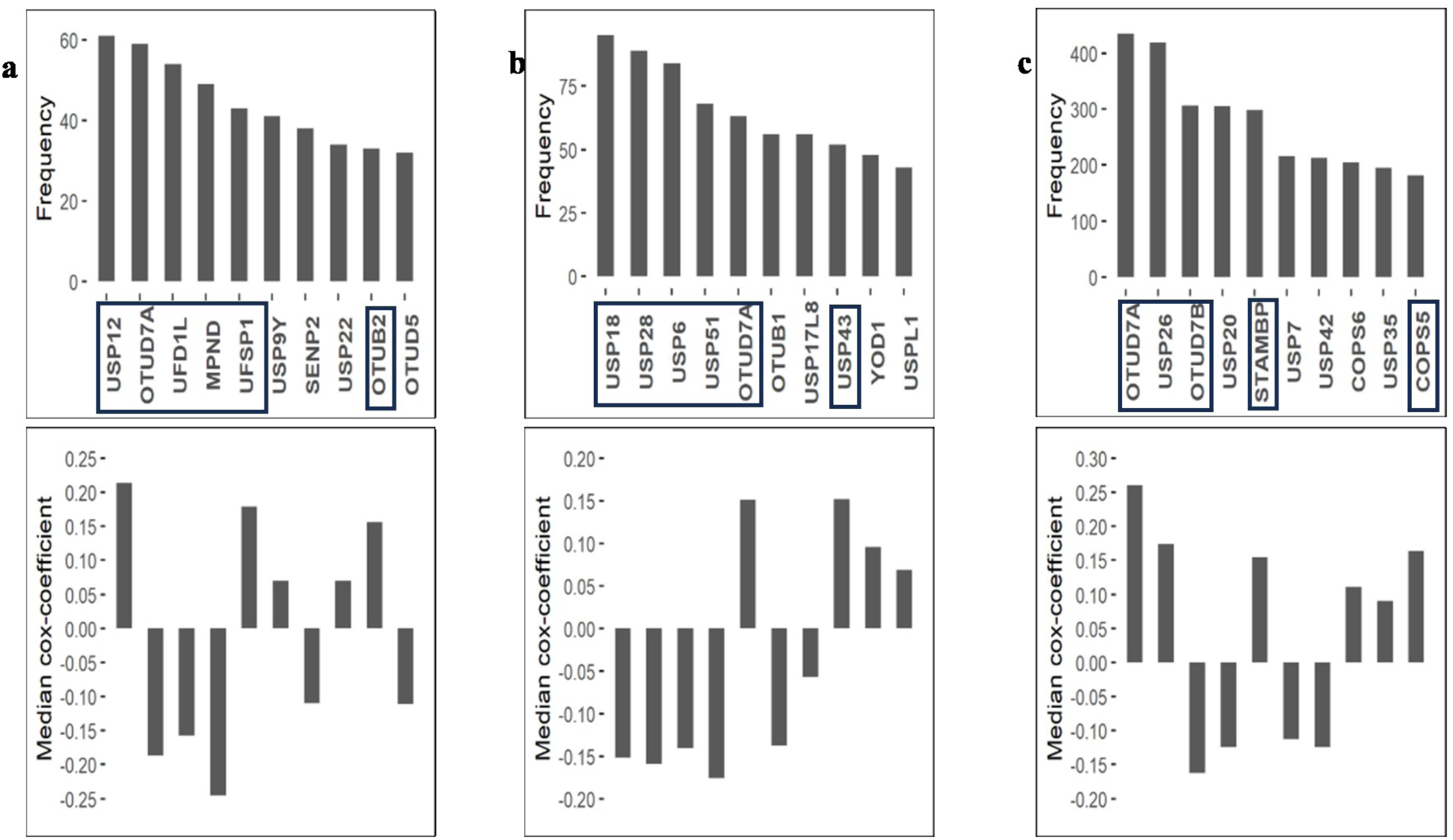
Top 10 most occurred DUBs in Cervical(a), Ovarian(b), and Uterine(c) cancers. Upper panel: Bar plots showing the number of times significant models picked up the DUB in 1000 trials. Lower panel: Median cox-coefficient of top 10 DUBs. Selected prognostic DUBs for DAS are represented with a box. DUB, Deubiquitinase; DAS, Deubiquitinase Associated Signature

**Figure S3:**
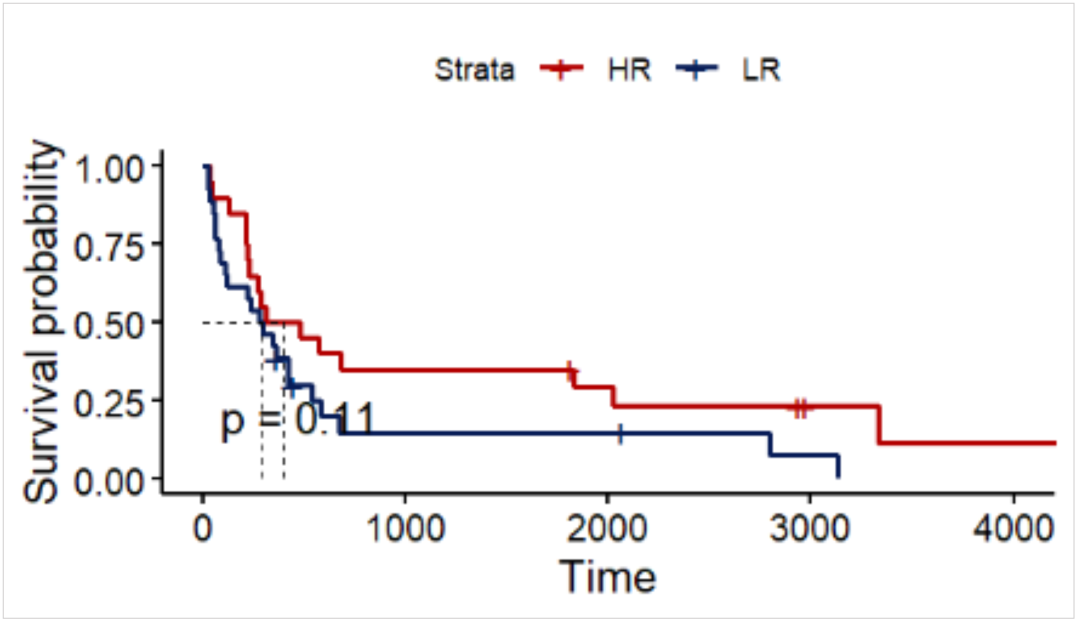
Validation of DAS in GEO dataset in Uterine cancer DAS, Deubiquitinase Associated Signature; GEO, Gene Expression Omnibus

**Figure S4:**
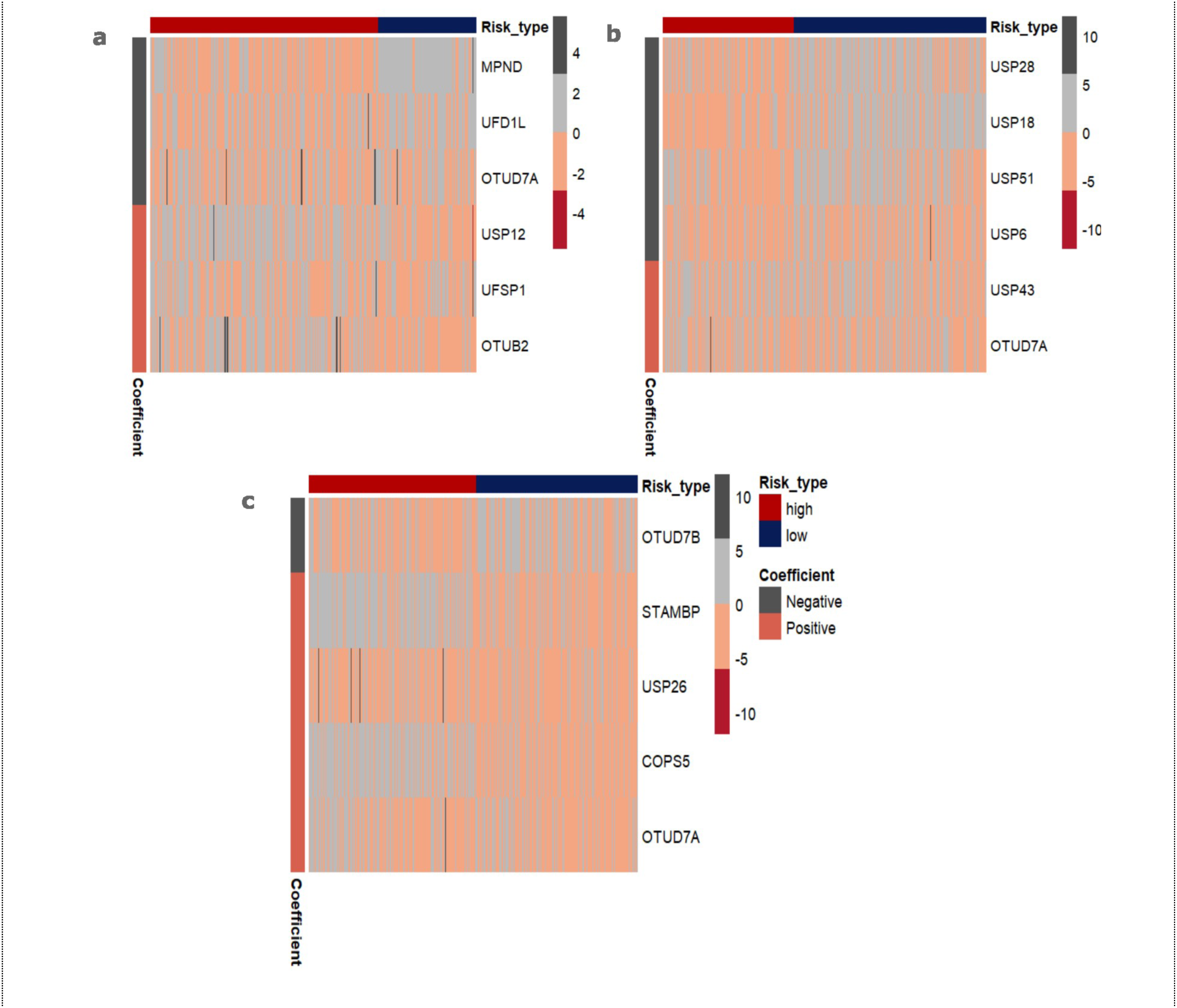
Heatmaps showing the expression of prognostic DUBs in Cervical(a), Ovarian(b) and Uterine(c) cancers. DUBs, Deubiquitinases

**Figure S5:**
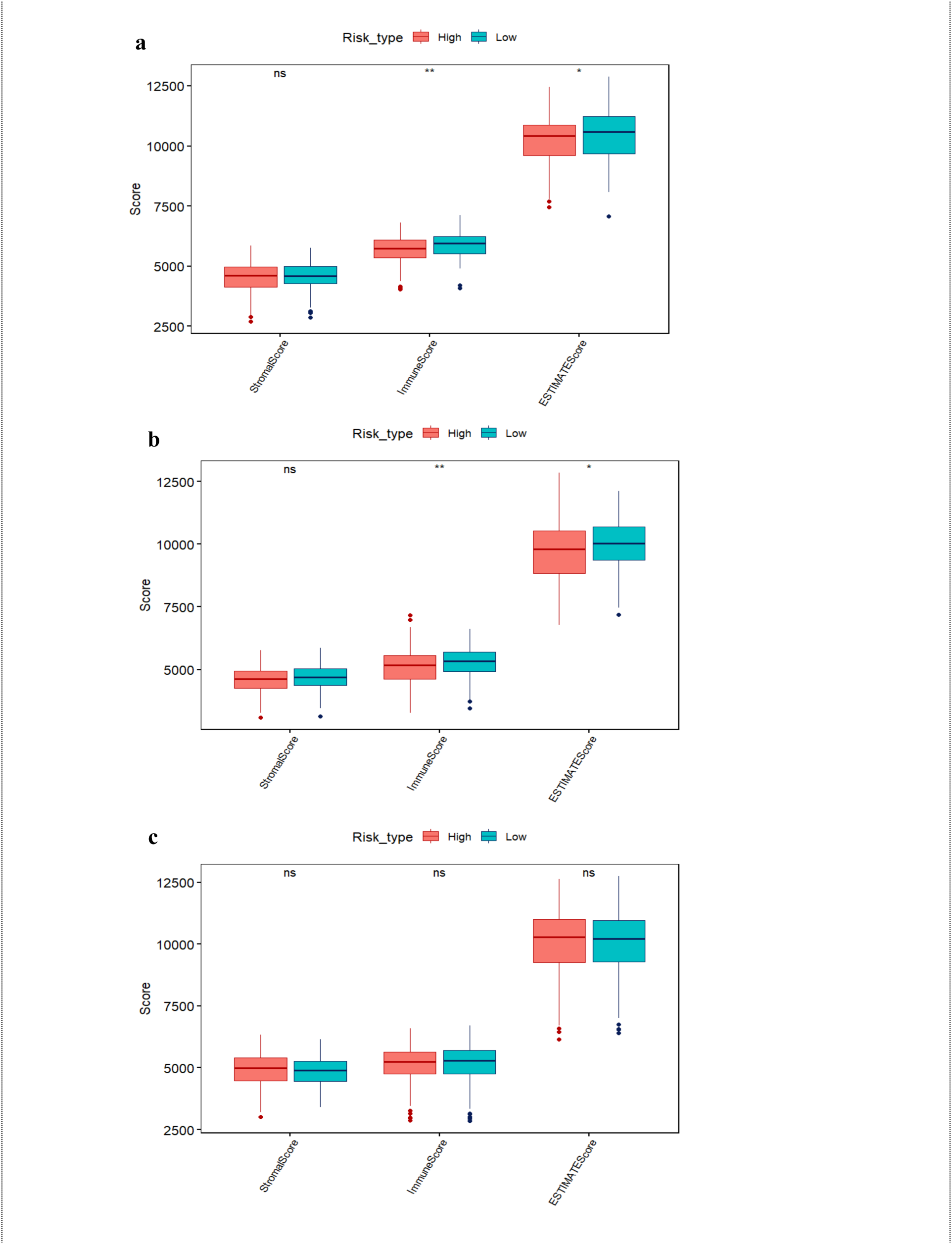
ESTIMATE scores of high-risk and low patients in Cervical(a), Uterine(b), and Ovarian cancers(c).

**Figure S6:**
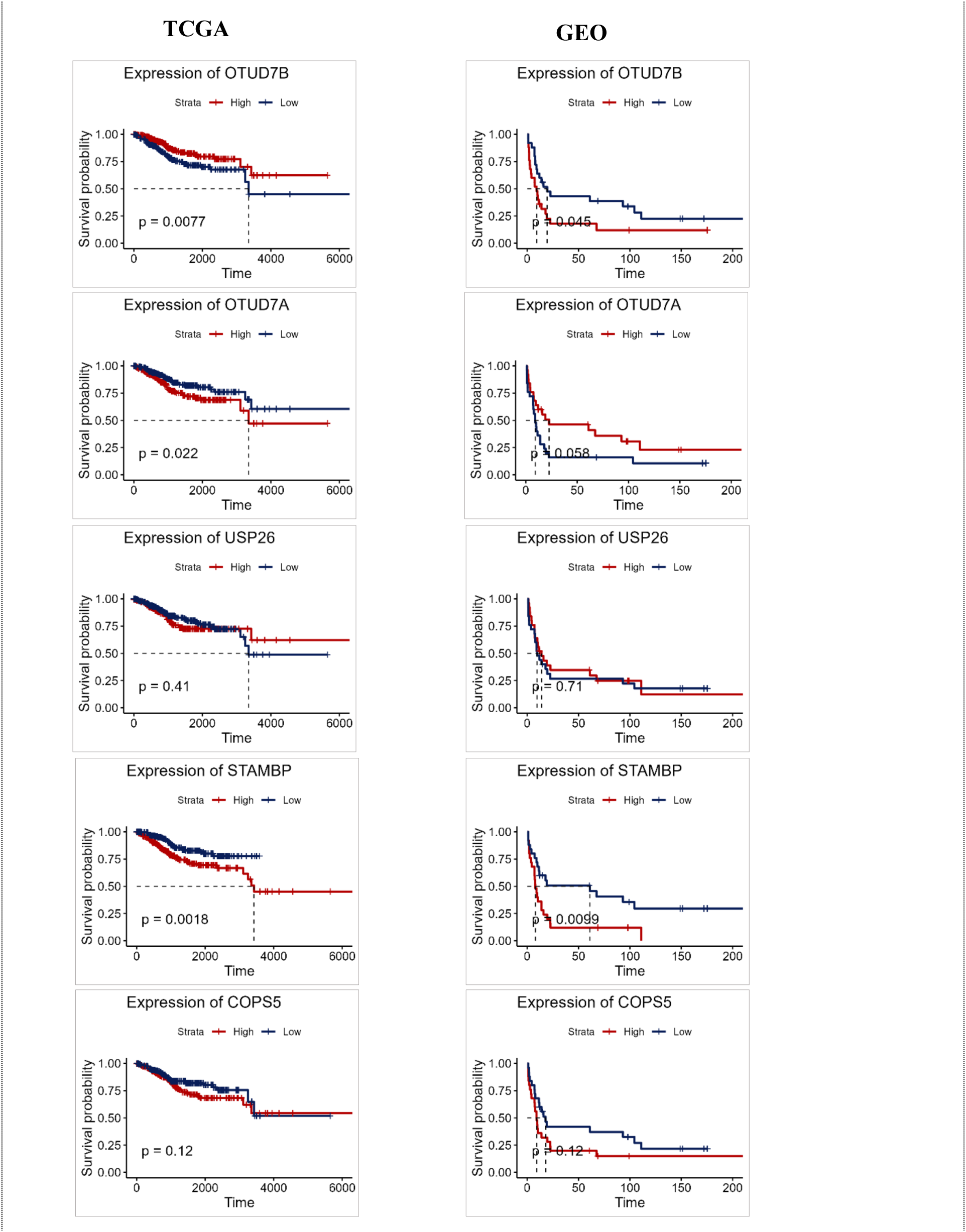
KM plots showing the relation between two risk groups and expression of prognostic DUBs in TCGA and GEO datasets of Uterine cancer. KM, Kaplan Meier; DUBs, Deubiqutinases; GEO, Gene Expression Omnibus; TCGA, The Cancer Genome Atlas

**Figure S7:**
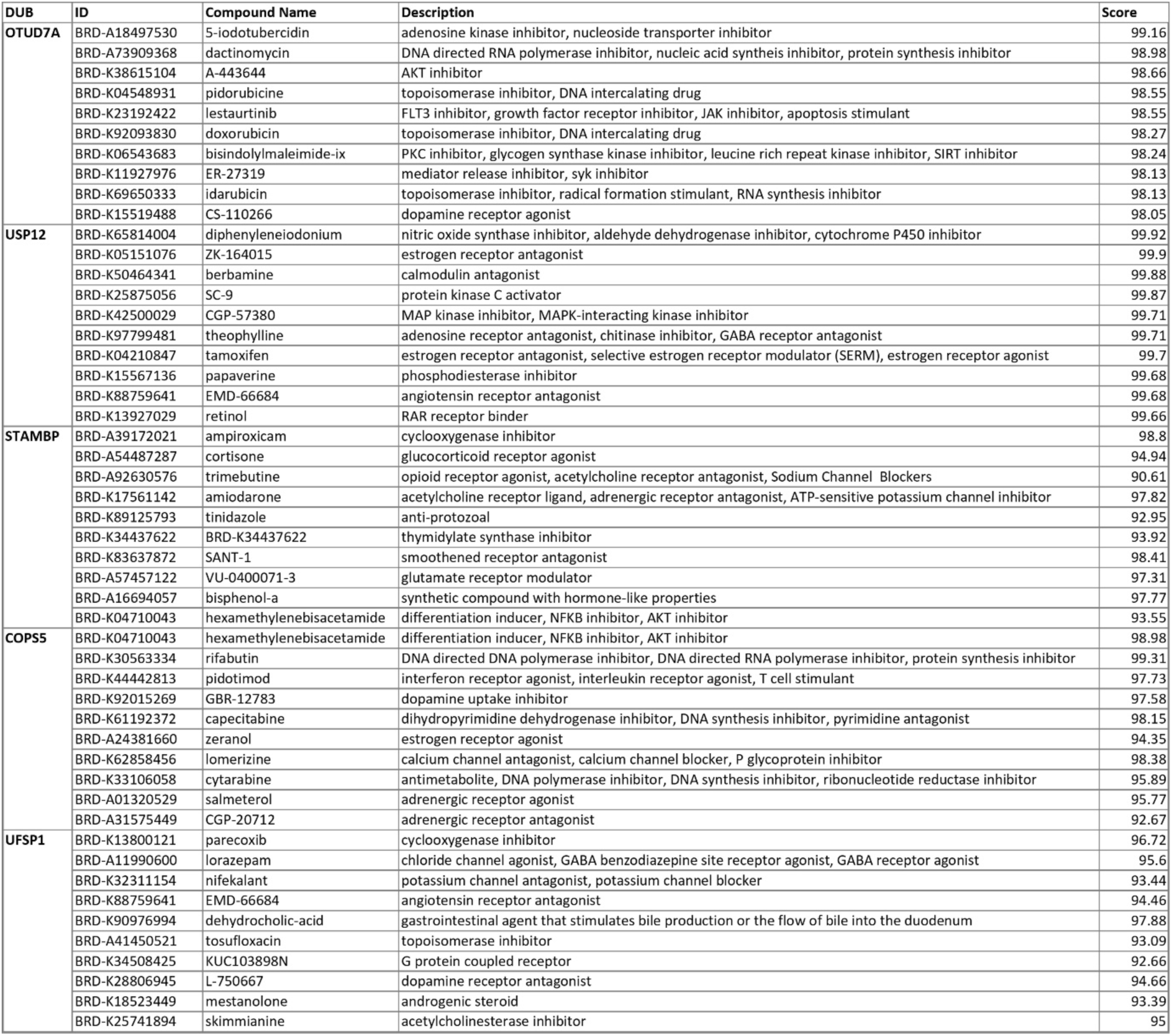
Result of CMap analysis for unfavorable DUBs CMap, Connectivity Map; DUBs, Deubiquitinases

**Table S1:** Sample information of TCGA and GEO datasets of three cancer types.

| Clinical features |  | Cervical Cancer |  | Ovarian Cancer |  | Uterine Cancer |  |
| --- | --- | --- | --- | --- | --- | --- | --- |
|  |  | TCGA-CESC | GSE44001 | TCGA-OV | GSE63885 | TCGA-UCEC | GSE119041 |
| Sample Size |  | 306 | 300 | 429 | 101 | 577 | 50 |
| Complete clinical information |  | 290 | 300 | 365 | 75 | 423 | 50 |
| OS | 0 | 218 | 262 | 140 | 5 | 352 | 11 |
|  | 1 | 72 | 38 | 225 | 70 | 71 | 39 |
| Grade | I | 18 | - | 1 | - | 98 | - |
|  | II | 135 | - | 42 | - | 120 | - |
|  | III | 118 | - | 320 | - | 318 | - |
|  | IV | 1 | - | 1 | - | 0 | - |
| MPS | Pre | 182 | - | 79 | - | 63 | - |
|  | Post | 106 | - | 283 | - | 463 | - |

## References

1. Siegel Mph RL, Miller KD, Sandeep N, et al. Cancer statistics, 2023. Wiley Online Library. 2023;73(1):17–48. doi:10.3322/caac.21763

2. Zhu W, Xie L, Han J, Cancers XG, 2020 undefined. The application of deep learning in cancer prognosis prediction. mdpi.comW Zhu, L Xie, J Han, X GuoCancers, 2020•mdpi.com. Accessed March 5, 2025. https://www.mdpi.com/2072-6694/12/3/603

3. Liu W, Jiang Q, Sun C, et al. Developing a 5-gene prognostic signature for cervical cancer by integrating mRNA and copy number variations. SpringerW Liu, Q Jiang, C Sun, SH Liu, Z Zhao, D WuBMC cancer, 2022•Springer. 2021;22(1). doi:10.1186/s12885-022-09291-z

4. Qu X, Shi Z, Guo J, et al. Identification of a novel six-gene signature with potential prognostic and therapeutic value in cervical cancer. Wiley Online Library. 2021;10(19):6881–6896. doi:10.1002/cam4.4054

5. Yuan L, Lu Z, Sun G, Medicine DC, 2022 undefined. Identification and verification of a 4-gene signature predicting the overall survival of cervical cancer. *journals.lww.com*. Accessed March 5, 2025. https://journals.lww.com/md-journal/fulltext/2022/10210/Identification_and_verification_of_a_4_gene.5.aspx

6. Pan X, Chen Y, Gao S. Four genes relevant to pathological grade and prognosis in ovarian cancer. Cancer Biomarkers. 2020;29(2):169–178. doi:10.3233/CBM-191162

7. Yang L, Jing J, Sun L, Medicine YY, 2018 undefined. Exploring prognostic genes in ovarian cancer stage-related coexpression network modules. journals.lww.comL Yang, J Jing, L Sun, Y YueMedicine, 2018•journals.lww.com. Accessed March 5, 2025. https://journals.lww.com/md-journal/fulltext/2018/08240/Exploring_prognostic_genes_in_ovarian_cancer.42.aspx

8. Liu J, Feng M, Li S, et al. Identification of molecular markers associated with the progression and prognosis of endometrial cancer: a bioinformatic study. SpringerJH Liu, M Feng, SY Li, S Nie, H Wang, S Wu, J Qiu, J Zhang, WJ ChengCancer cell internaOonal, 2020•Springer. 2020;20(1). doi:10.1186/s12935-020-1140-3

9. Yu SH, Cai JH, Chen DL, et al. LASSO and bioinformatics analysis in the identification of key genes for prognostic genes of gynecologic cancer. mdpi.comSH Yu, JH Cai, DL Chen, SH Liao, YZ Lin, YT Chung, JJP Tsai, CCN WangJournal of personalized medicine, 2021•mdpi.com. Published online 2021. doi:10.3390/jpm11111177

10. Li Y, Chen Z, Wu Y, et al. Unraveling role of ubiquitination in drug resistance of gynecological cancer. pmc.ncbi.nlm.nih.govL Yu, Z Chen, Y Wu, M Xu, D Zhong, H Xu, W ZhuAmerican Journal of Cancer Research, 2024•pmc.ncbi.nlm.nih.gov. 2024;14(5):2523–2537. doi:10.62347/WYKZ9784

11. Liu F, Chen J, Li K, et al. Ubiquitination and deubiquitination in cancer: from mechanisms to novel therapeutic approaches. SpringerF Liu, J Chen, K Li, H Li, Y Zhu, Y Zhai, B Lu, Y Fan, Z Liu, X Chen, X Jia, Z Dong, K LiuMolecular Cancer, 2024•Springer. 2024;23(1). doi:10.1186/s12943-024-02046-3

12. Hao Y, Muloye Guy M, Liu Q, et al. Construction of a prognostic model based on eight ubiquitination-related genes via machine learning and potential therapeutics analysis for cervical cancer. fronOersin.orgY Hao, MM Guy, Q Liu, R Li, Z Mao, N Jiang, B Wang, B Cui, W ZhangFronOers in GeneOcs, 2023•fronOersin.org. 2023;14:1142938. doi:10.3389/fgene.2023.1142938

13. Fernandez-Retana J, … HZMT, 2017 undefined. Gene signature based on degradome-related genes can predict distal metastasis in cervical cancer patients. journals.sagepub.comJ Fernandez-Retana, H Zamudio-Meza, M Rodriguez-Morales, A Pedroza-TorresTumor Biology, 2017•journals.sagepub.com. 2017;39(6). doi:10.1177/1010428317711895

14. Yan Z, Yu J, Wang S, et al. Identification of E3 ubiquitin ligase-associated prognostic genes and construction of a prediction model for uterine cervical cancer based on bioinformatics analysis. SpringerZ Yan, J Yu, S Wang, W Wen, M Xin, X LiDiscover Oncology, 2024•Springer. 2024;15(1):395. doi:10.1007/s12672-024-01271-y

15. Feng Y, Shan L, Gong Y, et al. Identification of ubiquitin markers for survival and prognosis of ovarian cancer. cell.comY Feng, L Shan, Y Gong, W Hang, Z Sang, Y Sun, K Tang, Y Wang, B Hu, X XiHeliyon, 2024•cell.com. Accessed March 5, 2025. https://www.cell.com/heliyon/fulltext/S2405-8440(24)13319-1?uuid=uuid%3A7ad0ed33-e5a3-4082-805b-f0364db33434

16. Topno R, Singh I, Kumar M, cancer PAB, 2021 undefined. Integrated bioinformatic analysis identifies UBE2Q1 as a potential prognostic marker for high grade serous ovarian cancer. SpringerR Topno, I Singh, M Kumar, P AgarwalBMC cancer, 2021•Springer. 2021;21(1). doi:10.1186/s12885-021-07928-z

17. Luo D, Li X, Wei L, et al. Ubiquitin-related gene markers predict immunotherapy response and prognosis in patients with epithelial ovarian carcinoma. nature.comD Luo, X Li, L Wei, Y Yu, Y Hazaisihan, L Tao, S Li, W JiaScienOfic Reports, 2024•nature.com. doi:10.1038/s41598-024-76945-2

18. Wang Z, Cheng S, Liu Y, et al. Gene signature and prognostic value of ubiquitination-related genes in endometrial cancer. SpringerZ Wang, S Cheng, Y Liu, R Zhao, J Zhang, X Zhou, W Shu, D Feng, H WangWorld journal of surgical oncology, 2023•Springer. 2023;21(1):3. doi:10.1186/s12957-022-02875-w

19. Zhang J, Wu G, Peng R, et al. A novel scoring model of deubiquitination paVerns predicts prognosis and immunotherapeutic response in hepatocellular carcinoma. ElsevierJ Zhang, G Wu, R Peng, J Cao, D Tu, J Zhou, B Su, S Jin, G Jiang, C Zhang, D BaiTranslaOonal Oncology, 2023•Elsevier. Accessed March 5, 2025. https://www.sciencedirect.com/science/article/pii/S1936523323001754

20. Chatrath A, Przanowska R, Kiran S, et al. The pan-cancer landscape of prognostic germline variants in 10,582 patients. Genome Med. 2020;12(1). doi:10.1186/s13073-020-0718-7

21. Kosary CL. Figo stage, histology, histologic grade, age and race as prognostic factors in determining survival for cancers of the female gynecological system: An analysis of 1973-87 SEER cases of cancers of the endometrium, cervix, ovary, vulva, and vagina. Semin Surg Oncol. 1994;10(1). doi:10.1002/ssu.2980100107

22. Xie L, Chu R, Wang K, et al. Prognostic Assessment of Cervical Cancer Patients by Clinical Staging and Surgical-Pathological Factor: A Support Vector Machine-Based Approach. Front Oncol. 2020;10. doi:10.3389/fonc.2020.01353

23. Zhao M, Wen K, Fan X, et al. OTUD7A Regulates Inflammation-and Immune-Related Gene Expression in Goose FaVy Liver. Agriculture (Switzerland). 2022;12(1). doi:10.3390/agriculture12010105

24. Su S, Chen J, Jiang Y, et al. SPOP and OTUD7A Control EWS–FLI1 Protein Stability to Govern Ewing Sarcoma Growth. Wiley Online LibraryS Su, J Chen, Y Jiang, Y Wang, T Vital, J Zhang, C Laggner, KT Nguyen, Z Zhu, AW PrevabeAdvanced Science, 2021•Wiley Online Library. 2021;8(14). doi:10.1002/advs.202004846

25. Zuo S, Wei M, Wang S, Dong J, Wei J. Pan-Cancer Analysis of Immune Cell Infiltration Identifies a Prognostic Immune-Cell Characteristic Score (ICCS) in Lung Adenocarcinoma. Front Immunol. 2020;11. doi:10.3389/fimmu.2020.01218

26. Dessources K, Ferrando L, Zhou QC, et al. Impact of immune infiltration signatures on prognosis in endometrial carcinoma is dependent on the underlying molecular subtype. Gynecol Oncol. 2023;171. doi:10.1016/j.ygyno.2023.01.037

27. Zhou X, Zhao Y, Ling ZJ, Yang B. Profiles of immune infiltration in ovarian cancer and their clinical significance: A gene expression-based study. Eur J Gynaecol Oncol. 2021;42(2). doi:10.31083/j.ejgo.2021.02.5347

28. Truxova I, Kasikova L, Hensler M, et al. Mature dendritic cells correlate with favorable immune infiltrate and improved prognosis in ovarian carcinoma patients. J Immunother Cancer. 2018;6(1):139. doi:10.1186/S40425-018-0446-3

29. Nersesian S, Glazebrook H, Toulany J, Grantham SR, Boudreau JE. Naturally Killing the Silent Killer: NK Cell-Based Immunotherapy for Ovarian Cancer. Front Immunol. 2019;10. doi:10.3389/fimmu.2019.01782

30. Zhang A, Zhang L, Xie X, Liu D. Inhibition of ATM with KU-55933 Sensitizes Endometrial Cancer Cell Lines to Olaparib. Onco Targets Ther. 2023;16:1061. doi:10.2147/OTT.S426923

31. Liefers-Visser JAL, Meijering RAM, Reyners AKL, van der Zee AGJ, de Jong S. IGF system targeted therapy: Therapeutic opportunities for ovarian cancer. Cancer Treat Rev. 2017;60. doi:10.1016/j.ctrv.2017.08.012

32. Jacome Sanz D, Raivola J, Karvonen H, et al. Evaluating targeted therapies in ovarian cancer metabolism: Novel role for pcsk9 and second generation mtor inhibitors. Cancers (Basel). 2021;13(15). doi:10.3390/cancers13153727

33. Ferdous J, Khatun S. Menopause and Gynecological Malignancy. Journal of SAFOMS. 2013;1(2). doi:10.5005/jp-journals-10032-1017

34. Spandidos DA, Dokianakis DN, Kallergi G, Aggelakis E. Molecular basis of gynecological cancer. In: Annals of the New York Academy of Sciences. Vol 900. ; 2000. doi:10.1111/j.1749-6632.2000.tb06216.x

35. Suga S, Kato K, Ohgami T, et al. An inhibitory effect on cell proliferation by blockage of the MAPK/estrogen receptor/MDM2 signal pathway in gynecologic cancer. Gynecol Oncol. 2007;105(2). doi:10.1016/j.ygyno.2006.12.030

36. McMellen A, Woodruff ER, Corr BR, Bitler BG, Moroney MR. Wnt signaling in gynecologic malignancies. Int J Mol Sci. 2020;21(12). doi:10.3390/ijms21124272

37. Tang LJ, Li Y, Liu YL, Wang JM, Liu DW, Tian QB. USP12 regulates cell cycle progression by involving c-Myc, cyclin D2 and BMI-1. Gene. 2016;578(1). doi:10.1016/j.gene.2015.12.006

38. Xiao J, Wang L, Zhuang Y, Zhu Q, … WLAJ of, 2024 undefined. The deubiquitinase OTUB2 promotes cervical cancer growth through stabilizing FOXM1. ncbi.nlm.nih.gov. Accessed July 23, 2024. https://www.ncbi.nlm.nih.gov/pmc/articles/PMC10839374/

39. Song Y, Wu Q. RBM15 m6A modification-mediated OTUB2 upregulation promotes cervical cancer progression via the AKT/mTOR signaling. Environ Toxicol. 2023;38(9). doi:10.1002/tox.23852

40. Pei L, Zhao F, Zhang Y. USP43 impairs cisplatin sensitivity in epithelial ovarian cancer through HDAC2-dependent regulation of Wnt/β-catenin signaling pathway. Apoptosis. 2024;29(1-2). doi:10.1007/s10495-023-01873-x

